# “Rites of Passage: Professional Identity Formation and the OTOHNS Oral Board Exam”

**DOI:** 10.64898/2026.03.19.26347858

**Authors:** Kevin C. McMains

**Author notes:** Corresponding Author: Kevin C. McMains, MD, 7400 Merton Minter Dr, San Antonio, TX 78229. Funding: American Board of Medical Specialties Visiting Scholars Grant. COI: None.

## Abstract

**Objectives:** Professional Identity Formation has been defined as an individual internalizing the values and norms of the medical profession in ways that result in thinking, acting, and feeling like a physician. During the COVID-19 Pandemic, the ABOHNS pivoted the format of the oral board exam from in-person exams to virtually administered exams. In light of this, we ask:

I. How, if at all, do Otolaryngology-Head and Neck Surgery Oral Board Examinations shape examinee professional identity?
II. Do different formats of administering Otolaryngology-Head and Neck Surgery Oral Board Examinations have different effects on examinee Professional Identity Formation (PIF)?

**Methods:** Thematic analysis was used to explore candidate experience. We developed and tested a shortened Professional Identity Essay that foregrounds the PIF effects resulting from differing methods of administering the Oral Board Examination. Themes generated from semi-structured interviews were compared to identify differences Professional Identity resulting from OBEs.

**Results:** Nineteen participants enrolled in our study, each completing a single interview lasting between 15-30 minutes. We found participants responses to coalesce around 3 themes: educational effect of the OBE on PIF; different OBE formats carried distinct stresses; and the catalytic effect on PIF from in-person OBE.

**Conclusions:** Participating in either format of the ABOHNS OBE demonstrated and educational effect on PIF. Additionally, when delivered in an in-person format, the ABOHNS OBE also catalyzed ongoing PIF. This effect of the OBE offers an additional potent mechanism to integrate the most inclusive range of candidates into the community of Otolaryngology practice.

**Level of Evidence: VI:** (Single Qualitative Study investigating perspectives of healthcare providers on a specific intervention)

## Introduction

One of the characteristics of Professions is a contract with society.^1^ It is in service of this contract that the American Board of Otolaryngology-Head and Neck Surgery (ABOHNS) Board conducts written qualifying and oral certifying examinations for those who meet eligibility requirements. At the post-graduate education level, the ACGME articulated core competencies to guide desired outcomes of medical trainees. Professionalism is one of these competencies. While professionalism can be taught, Hafferty observes that what the profession should be concerned with is “What one *is* rather than what one *does*”.^2^ Said differently, the concern goes beyond external, observable actions of professionalism towards something deeper: development of professional identity.

Since the Carnegie Report focused attention on the importance of Professional Identity Formation (PIF) in medical education, PIF has become of central interest to design and implementation of medical education.^3^ PIF has been defined as an individual internalizing the values and norms of the medical profession in ways that result in thinking, acting, and feeling like a physician.^4^ In addition to attesting to clinical reasoning, one of the primary aims of GME training and of Board certification should be to support individuals as they develop their professional identities.^4^ Let us consider the process of identity development more broadly. Erikson described sequential steps of psychological development over a life cycle and theorized that, in order to move from one stage to the next, an individual had to undergo a crisis of identity.^5^ At the transition from one developmental stage to another, the individual’s ideas and actions meet a challenge that requires development of new capabilities and different understanding of self. Kegan argued that the concept of identity development also applies to the professional setting.^6^ Consistent with this developmental model, Cantillon et al demonstrated progressive identification with a trainee’s chosen specialty as a result of post-graduate training.^7^ Additionally, specific “rites of passage”, with their embedded symbols and rituals, aid and encourage significant life transitions in part through situating the identity crisis experienced within a community context.^8^ Given the high-stakes nature of oral board exams, it is conceivable that participating in the oral board examination could serve as a “rite of passage”, catalyzing a watershed moment of PIF.

During the COVID-19 Pandemic, the ABOHNS pivoted the format of the oral board exam to deliver on the social contract in a safe, defensible manner. There are several criteria of effective programs of assessment.^9^ One of the criteria is the assessment’s catalytic effect, which is to say how it affects the learner’s trajectory of growth following the assessment. While the enforced change in format resulted in a psychometrically-valid examination, the question of whether learners gain the valuable experience of a rite of passage remains unanswered.

How to assess downstream effects of the OBE on PIF represents a complex challenge. Leveraging reflective practice provides a potential solution to this problem. Reflective practice can contribute to professional identity formation development of professional values.^10,11^ One tool for this is Bebeau’s “Professional Identity Essay”, a set of open-ended questions designed to elicit the respondent’s conception of the professional’s role in society.^1^ Learners’ responses to this prompt offer a valuable window into PIF.

With these background considerations in mind, we frame these research questions: How, if at all, do Otolaryngology-Head and Neck Surgery Oral Board Examinations shape examinee professional identity? Do different formats of administering Otolaryngology-Head and Neck Surgery Oral Board Examinations have different effects on examinee professional identity formation?

## Methods

In this study, thematic analysis was used for a comprehensive analysis of candidate experience. We developed and tested an interview guide based on the Professional Identity Essay that emphasizes the PIF effects resulting from differing methods of administering the OBE. (Appendix) Themes generated from semi-structured interviews were compared to explore differences Professional Identity resulting from OBEs. The study was conducted in a manner consistent with the Standards for Reporting Qualitative Research (SRQR) checklist.^12^

### Context

In this qualitative interview study, we explored Otolaryngologists’ trajectories of PIF by sampling practicing Otolaryngology graduates from United States Graduate Medical Education (GME) programs who participated in the last year of in person OBEs and the first year of virtual OBEs.

### Sampling strategy

All Otolaryngologists who completed residency training in 2019 and 2020 and sat for the OTOHNS OBE were invited to participate. We chose this group for our sample to map trajectory of PIF since OBE. Additionally, the different format of OBE experienced by each cohort provided a natural control group. Participants were recruited by email from the ABOHNS. The email was sent twice to each eligible Otolaryngologist by a Board staff member who had no direct involvement with certification decisions.

### Data collection

Semi-structured interviews were conducted by the research assistant using an interview guide exploring PIF, using the PIE as a foundation. (Appendix) The interviews were conducted between November 2023 and April 2024, each lasting between 15-30 minutes. Interviews focused on participants’ lived experiences surrounding the OBE and how these experiences shaped their PIF. Interviews were recorded, transcribed, checked for accuracy, and de-identified. Specific demographic identifiers such as gender were redacted to the degree possible to assure anonymity.

### Data analysis

Coding and thematic analysis were conducted iteratively by the author. The interviews offered a rich and varied picture of the experiences of these physicians, forming a sufficient data source to answer our research questions.^13^

### Techniques to Enhance Trustworthiness

As transcripts were coded transcripts, memos were maintained, data were interpreted and further revised to ultimately define themes. The final codes drawn from inductive coding, were: 1. Educational effect of OBE on PIF; 2. Different formats lead to different stresses; 3. Rite of Passage: Catalytic Effect on PIF from in Person OBE. These themes brought into focus the characteristics, values, and norms our participants drew upon as they worked to think, act, and feel like Otolaryngologists at the threshold of their professional careers. This study was approved by the XXX University IRB (Reference number: 1730820-1)

### Researcher Characteristics and Reflexivity

KCM is a clinician educator and PhD candidate. The author brings privilege as a white, male educator and considered this explicitly to better understand how participants’ social positions affected their trajectories.

## Results

Nineteen participants enrolled in our study, each completing a single interview lasting under between 15-30 minutes. Of these, 9 participated in the In-Person OBE format and 10 participated in the Virtual OBE format. In order to preserve anonymity, individual demographic data were not collected and pseudonyms were created for participant where quoted.

For the research questions under investigation, we found participants responses to coalesce around 3 themes: educational effect of the OBE on PIF; different OBE formats carried distinct stresses; and the catalytic effect on PIF from in-person OBE.

### Educational Effect of OBE on PIF

For examinees who underwent both formats of OBE, they described significant IF during the course of residency training. Neither group described significant effect on PIF resulting from preparation for the written (qualifying) exam. By contrast, both groups narrated significant PIF resulting from the process of preparing for the OBE. (Table 1) Taken together, these data reflect that there is an educational effect from the OBE that crystalizes the PIF underway throughout residency and is distinct from the PIF effects resulting from the written/qualifying exam.

### Different OBE formats carried distinct stresses

Participants described distinct stresses attendant to the OBE format in which they participated. The prospect of an IP exam carried an increased emotional load. (Table 1) Morgan elaborated “virtual was just so convenient. I didn’t have to pay for any flights. I didn’t have to pay for a hotel. I didn’t have to fly out anywhere.”

While travel considerations were not significant for the V format, uncertainty surrounding technology suffused the experience of this group. Finally, some female participants (e.g., those who talked about their gender explicitly) talked about how the V format allowed them to take part in the exam. Amrit confided

> “I was actually 38 weeks pregnant when I took my oral boards…I was able to, you know, take my exam at a time when, due to pregnancy, I wouldn’t have been cleared to travel, so I was able to kind of like complete that step in my professional journey, when otherwise it would have probably been delayed.”

In this way, for some participants, the virtual format provided an avenue to continue the certification process that would have otherwise been delayed due to pregnancy.

### Rite of Passage: Catalytic Effect on PIF from In-Person OBE

While both V and IP groups described PIF resulting from *preparation for* the OBE (educational effect), only participants in the IP group consistently described PIF being shaped by *participation in* the OBE. The IP group described a greater sense of connection to the Otolaryngology Community of Practice (COP), both with other examinees and the examiners. (Table 1)

This difference in experience led many within the V group to question the legitimacy of their OBE experience. (Table 1) These impressions contrast with Campbell’s (IP) refection on that: *“I walked out of there and I was like ‘I did a great job*.*’ And I felt really, it made me feel a lot more confident in the fact that I could practice and take care of people*.*”*

The cumulative effect of the difference in sense of connection to the community of Otolaryngologists and lingering doubts about the legitimacy of the exam format resulted in distinct trajectories of PIF between V and IP groups. Campbell (IP) expresses a change in self-efficacy resulting from the OBE:

> *“And there’s been a continued evolution of my professional identity over the course of the five years that I’ve been doing this since finishing fellowship. But I don’t, I can’t really imagine having started practice before taking that test. I felt like that it was like a stamp. ‘Like you can do this. You’re allowed to do this, and you know how to do this*.*’”*

Overall, participants in the IP group felt increased connection to the Otolaryngology COP as well as a greater sense of legitimacy stemming from their OBE experience. As a result, IP examinees’ OBE experience set them on a distinct PIF trajectory from the V group.

## Discussion

To our knowledge, this was the first deliberate exploration of the effect of the Oral Board (Certifying) Exam on PIF among graduates from Otolaryngology GME programs. We were able to explore the effects on PIF resulting from preparation for and participation in the OBE. Additionally, interviewing OBE examinees who underwent the OBE in different formats allowed comparison of PIF effects between groups. Three themes were identified from our dataset:

*Educational Effect of OBE on PIF; Different OBE formats carried distinct stresses; Rite of Passage: Catalytic Effect on PIF from In-Person OBE*.

A few words about what constitutes trustworthy assessment: In 2018 the Ottawa Conference convened a group to determine criteria by which systems of assessments in medical education should be evaluated. This group produced the 2018 Consensus framework for good assessment.^9^ The criteria proposed included validity, reproducibility, equivalence, feasibility, educational effect, catalytic effect, and acceptability. While some criteria are viewed as more important for formative assessment (validity, educational effect, catalytic effect), others are regarded as essential for summative assessments (e.g. validity, reproducibility, equivalence).

Appropriate to a process aimed at preserving the public trust, Board Certification emphasizes characteristics of summative assessment. The mission statement of the ABOHNS reads: “The American Board of Otolaryngology - Head and Neck Surgery serves the public by assuring that diplomates meet our standards of training, knowledge and professionalism through initial and continuing certification.”^14^ One of the stated objectives of the ABOHNS is “To ensure those who attain board certification continue to meet the standards set forth by the ABOHNS throughout their careers.” Initial board certification in OTOHNS occurs in two stages-the written Qualifying Exam and the Oral Certifying Exam. Once initially certified, all diplomates of the ABOHNS are required to participate in an ongoing Maintenance of Certification (MOC) process. Together, these represent a system of assessment. While examinations leading to Board certification are traditionally considered summative assessments,^9^ the ongoing nature of and need for MOC highlight the additional value of formative elements in certification. The data presented below strongly suggest formative impact resulting from the OBE.

### Educational Effect of OBE on PIF

Participants in neither cohort experienced significant influence on PIF from preparing for the written certifying exam, citing similarity to other examinations taken throughout their academic careers. However, both groups reported significant PIF resulting from the process of preparing for the OBE. Miller described a pyramid of assessment-knows, knows how, show how, does.^15^ Among these levels, there is an inverse relationship between reproducibility and authenticity. The written certifying exam is an example of a knowledge exam, characterized by high levels of reproducibility, but abstracted form the clinical setting. By contrast, the OBE includes elements of the first 3 levels-knows, knows how (e.g. the candidate describes what they might do in a given situation), shows how (e.g. the candidate draws the incisions for a rotational flap to close a Moh’s defect). Of these forms of assessment, theory would predict OBE to have a greater effect on PIF. In their articulation of Situated Learning Theory, Lave and Wenger describe how PIF occurs as learners are allowed to participate in the authentic work of a Community of Practice (COP).^16^ In the case of the OBE, these data support sufficient authenticity of the examination to generate an educational effect on PIF resulting from preparing for the OBE. This process crystalizes the PIF begun during residency and continuing as they move into clinical practice.

### Different Formats Lead to Different Stresses

Participants in each method of OBE administration described distinct stresses arising from the experience. In-person participants cited travel-related stresses as well as a higher emotional load surrounding the face-to-face interaction. Virtual participants identified uncertainty around technology and the difficulty to find a location that met acceptable criteria as a testing space. Equity concerns were also raised about requiring IP examinations. Financial costs associated with travel and opportunity cost of lost days working may represent a burden that is unequally borne by candidates of different economic backgrounds and differing debt loads. The option of a virtual OBE allowed candidates in their 3^rd^ trimester of pregnancy to participate without delaying until the next examination cycle. Rowe, et al. catalogue the range of accommodations allowed by the 14 Boards requiring OBEs.^17^ They found a direct relationship between increased accommodations and female representation on Boards. The National Research Council affirms that accommodations can be provided during testing which give more accurate picture of the test-taker’s true abilities without compromising the validity of testing. Given these data and the many benefits accruing to the field of Otolaryngology from increased diversity,^18^ future iterations of the OBE should consider equity of access.

### Rite of Passage: Catalytic Effect on PIF from In-Person OBE

As organizations serving the public trust, certifying boards must assure that high standards of summative assessment are upheld. While different boards have approached OBEs in different ways, both in person and virtual OBEs demonstrate psychometric integrity.^17,19^ Despite psychometric equivalency between OBE formats, candidates who underwent the virtual exam questioned the legitimacy of their experience, leading to a form of imposter syndrome.^20^ This differed from those who underwent the OBE in the in-person format. For IP examinees, the OBE had a catalytic effect on PIF, representing an inflection point in their claim to identity as an Otolaryngologist. The OBE served symbolically as an initiation “rite of passage” for IP candidates. Blumenkrantz observes, “in contemporary American communities, few socially sanctioned, community-based rites of passage exist with enough breadth and depth to have an impact on an individual’s identity and sense of community.”^21^ The IP OBE can serve as an instrument of extending a sense of belonging within the profession to those who successfully complete it.^22^ Participants felt this sense of belonging extended not only to those with whom they sat the exam, but also to leaders in the field who administered it. To receive approval from these leaders cemented a sense of their own validity within the profession of Otolaryngology. Situated as it is within a trajectory of PIF development that begins in residency and continues through independent practice and MOC, participants in the in-person OBE experienced it as an initiation into Otolaryngology, crystallizing professional identity. As we consider both the *import* of ongoing PIF development beyond residency graduation, it is critical that the *impact* of the OBE on this process remains a focus.

### Limitations

There were several limitations to our study. Single interviews were conducted among members of a single clinical specialty within a North American setting. We acknowledge that Western contexts differ from those which may be present in Lower and Middle-Income countries.^23^ Also, given the fluidity of identity construction, a single interview cannot fully capture the developmental nature of PIF. Yet, the rich insight this population offers into developing identity as Otolaryngologists balance potential limits to transferability.

Additionally, the racial and gender characteristics of specific interviewees were not identified (unless explicitly discussed). This decision reflected a desire to ensure privacy of participants. Some richness of data may be lost as a result. Future work might specifically explore the experiences of people of color and women as they navigate PIF, while also negotiating their marginalized status.

## Conclusion

The ABOHNS serves the public trust by developing and administering examinations that lead to initial and ongoing Board Certification. While Board Certification has traditionally been considered a summative assessment, the suite of examinations within which the OBE is situated (written qualifying exam, oral certifying exam, and maintenance of certification), have characteristics of both formative and summative assessment. Participating in either format of the ABOHNS OBE demonstrated and educational effect on PIF. Additionally, when delivered in an in-person format, the ABOHNS OBE also catalyzed ongoing PIF. In light of this understanding, future OBE administration should be conducted in-person to capitalize on professional identity growth and sense of belonging within the profession experienced by successful applicants. This effect of the OBE offers an additional potent mechanism to integrate the most inclusive range of candidates into the community of Otolaryngology practice.

## Supporting information

Table: Rites of Passage

Appendix: Interview Guide

## Data Availability

All data produced in the present study are available upon reasonable request to the authors

## Acknowledgments

Grateful thanks to the American Board of Medical Specialties and the American Board of Otolaryngology/Head and Neck Surgery

