## Supplementary material for "“Rites of Passage: Professional Identity Formation and the OTOHNS Oral Board Exam”": Table: Rites of Passage

**Table 1: Themes and Additional Exemplar Quotes**

| Theme | Exemplar Quote |
| --- | --- |
| *Educational Effect of OBE on PIF* | *“I felt like this was what actually brought together the whole the whole package, the whole process of thinking, defending why you're thinking a certain way and even adding in like where you might deviate based on certain situations…to really put it together and go ‘I know this stuff and I know how to take care of it.’”* Campbell (IP) |
| *Different OBE formats carried distinct stresses* | Virtual  Technology  *“All of us in the back of our minds, you're just, like, worried about your Internet going out right and failing boards or something totally out of your control. So, I think that was an added, unusual source of stress from a virtual standpoint.” -Dakota (V)*  IP  Emotional Load  “As someone who is somewhat shy, I greatly appreciated doing it online as opposed to, you know, being feeling like I'm in front of someone and you know the in-person format.” -Riley (V)  Logistics  “You're so nervous and I think adding on to that like, you know, making sure that you're like, suitcase arrived on the plane with you.”  -Jesse (V)  Cost  “A lot of us are paying back student loans, a lot of us are paying back. We don't have the, at least at that time, didn't have the kind of money to be spending several hundred up to $1000 on a plane ticket and a room for several days and all that stuff.” -Jie (V) |
| *Rite of Passage: Catalytic Effect on PIF from In-Person OBE* | Community  “*I think there are huge advantages in my mind for community building and personal identity and social interactions when you're in person that all occur kind of around the main event.” -* Kim (IP)  *“I think there's a lot of camaraderie that goes along with going and doing it in person. And so I think when you take it virtually, you miss out on it on that. So you don't get to see people, the examiners and then also your sort of cohort that's taking it at the same time.” -Kai (V)* |
| *Questioning Legitimacy* | *“I do think that we weren't tested in the same way that they were. I don't think it's made me doubt, sort of, my training or anything, but I would probably say that my experience was not the same as theirs*.” -Kai (V)  “*“I honestly feel like I got off easy with the new format…. I think the in person had a lot more cases than the* ***actual*** *oral boards*.*”* (emphasis added) -Dakota (V) |

Key:

PIF- Professional Identity Formation

OBE- Oral Board Exam

IP- Underwent In-Person Oral Board Exam

V- Underwent Virtual Oral Board Exam
