## Appendix: Interview Guide for "“Rites of Passage: Professional Identity Formation and the OTOHNS Oral Board Exam”"

**Research Question:** How, if at all, does preparing for and participating in the Otolaryngology/Head and Neck Surgery Oral Board Exam (OBE) affect Professional Identity Formation?

**Sub-question is:**  Does the format of the Otolaryngology/Head and Neck Surgery Oral Board Exam (In-Person vs Virtual) affect Professional Identity Formation differently?

**Introduction:**

As we emerge from the COVID pandemic, the ABOHNS Board is considering how best to conduct oral board exams moving forward. Prior to the pandemic, the oral exam was conducted in person. The pandemic forced a pivot to conduct the oral exam remotely.

In executing the Board’s obligation to the public- assuring certification of competent Otolaryngologists- one dimension of competence that is important to explore is that of professional identity formation (PIF).

For the purpose of our discussion, we will define professional identity formation as: “An adaptive developmental process that happens simultaneously at two levels: (1) at the level of the individual, which involves the psychological development of the person and (2) at the collective level, which involves the socialization of the person into appropriate roles and forms of participation in the community’s work.” Or, more simply, the process by which a person comes to “think, act, and feel like a physician.”

We recognize that one potential impact of the Oral Board exam is that of a rite of passage, influencing a candidate’s professional identity journey. The ABOHNS is interested in your experience with the Oral Board Exam. Specifically, how, if at all, preparing for and participating in the Oral Board Exam in the format you participated in affected your professional identity journey?

Do you have any questions before we start?

Interview Questions for ABOHNS:

1. Tell me a story about what it means for you to “think, act, and feel” like an Otolaryngologist? Probe:
   a. Is there someone who you look to as an exemplar of an Otolaryngologist?
2. Tell me about what it was like preparing for your **written** board exam. Probes:
   a. How well did you feel your residency program prepared you?
   b. Did you prepare for the written exam exclusively individually or did you also prepare with a group?
   c. Did this preparation make you *feel* like an Otolaryngologist? In other words, did the preparation affect your professional identity as an Otolaryngologist in any noticeable way?
   d. And what about after? Did you feel “more” like an Otolaryngologist after, or did having taken the exam affect your Professional Identity?
3. Now tell me about what it was like preparing for your **oral** exam. Probes:
   a. How well did you feel like your residency program prepared you?
   b. Did you prepare for the Oral Board Exam exclusively individually? Or did you also prepare with a group?
   c. How did doing this preparation make you feel as an Otolaryngologist? In other words, how, if at all, was your professional identity affected by preparing for the oral exam?
   d. How did you feel after your oral exam? In particular, how did you feel as an Otolaryngologist? In other words, how, if at all, was your professional identity affected by participating in the oral exam?
4. We’ve talked about preparation for the Oral Board Exam. Now, tell me about what it was like **participating in** your **oral** exam. Probes:
   a. Did participating in the oral board exam make you *feel* like an Otolaryngologist? In other words, did participating in the Oral exam affect your professional identity as an Otolaryngologist in any noticeable way?
   b. And what about after? Did you feel “more” like an Otolaryngologist after, or how did having taken the oral exam affect your Professional Identity?
5. In what **format** did you participate? (in-person or virtual)? Probe:
   a. How, if at all, did the format of the Oral Board Exam affect your professional identity?
   b. How do you imagine things would have been different if you had participated in a different Oral Board Exam format? (in-person if your exam was virtual or virtual if your exam was in person)?
   c. Do you keep in touch/work with/etc anyone that took the other format? What have you noticed about others’ experiences or practices who had a different format than your own?
6. Would you please describe the type of **clinical practice** you have worked in since completing residency? (e.g. Academic, group private practice, hospital owned, solo, etc.) Probe:
   a. How have you felt/noticed your professional identity evolve since participating the Otolaryngology Oral Board Exam?
7. To conclude, is there anything that we discussed today that you feel should be emphasized or is of greater significance to you? Is there anything I missed related to professional identity formation or your experiences with the Otolaryngology Oral Board Exam that you would like to discuss?
